# Development of Untargeted Metabolomics-based Detection Method for *Naegleria fowleri*: A Year-Long Monitoring of the Microbial Ecology in an Operational Drinking Water Distribution System

**DOI:** 10.64898/2026.09.01.26358893

**Authors:** Zhihao Yu, Natalia Kulesza, Jason Wylie, Sergio Domingos, Brian H. Clowers, Geoffrey J. Puzon

## Abstract

*Naegleria fowleri* is known to be the causative agent of highly aggressive (> 95 % mortality rate) primary amoebic meningoencephalitis (PAM). *N. fowleri* can colonize drinking water distribution systems (DWDSs) which place additional pressures on water distribution authorities and public health officials. Chlorination and chloramination are widely adopted to combat *N. fowleri* in DWDSs but this approach is susceptible to failure in certain situations which makes effective *N. fowleri* surveillance necessary to protect the public from infection. Based on our previous efforts to develop a rapid *N. fowleri* detection method focusing on the lab-cultured and field-collected samples, this manuscript presents our recent progress investigating an operational DWDS field site seasonally colonized by *N. fowleri* on a monthly basis over the course of a year using untargeted metabolomics of the entire microbial ecology. A panel of significant features exhibiting changes between *N. fowleri* positive and negative samples were found and further correlated with the findings from the previous studies. The chemical identities for a subset of the common significant features were confirmed. A new, seasonal prediction model was built based on the common significant features and its performance was compared with our previous efforts. This study illustrates the further development of a rapid *N. fowleri* detection approach from a longitudinal perspective.

## 1. Introduction

*Naegleria fowleri* is a pathogenic free-living amoeba, known to be the causative agent of primary amoebic meningoencephalitis (PAM) that exhibits a fatality rate of 95%.(Visvesvara et al., 2007) The first report of *N. fowleri* came from Australia in 1965 and since then more than 400 cases have been reported globally across the United States, Australia and Pakistan, just to name a few.(Naqvi et al., 2016) The colonization sites of *N. fowleri* vary from natural systems (e.g. lakes, rivers, hot springs) (Barnhart et al., 2024; Shikany et al., 2025) and engineered systems (e.g. water parks and drinking water distribution systems (DWDSs)).(Cope et al., 2019, 2018; Malinowski et al., 2022, 2024; Miller et al., 2015; Morgan et al., 2016) The emergence of *N. fowleri* in DWDSs has caused multiple deaths in Australia, Pakistan, and in the United States (Arizona and Louisiana).(“Public Drinking Water Systems | Naegleria fowleri | CDC,” 2019) DWDSs can be seasonally colonised by *N. fowleri* and disinfection by chlorination is a widely adopted approach to fight against *N. fowleri*.(Puzon et al., 2020) For example, the Australian Government recommended 0.5 mg/L of free chlorine residual level throughout the water distribution systems.(Miller et al., 2017) In response to the findings of *N. fowleri* in parish water systems, the Louisiana Department of Health adopted the ‘2013 emergency rule’ where the disinfectant residual in the drinking water must maintain a continuous level of no less than 0.5 mg/L.(Miller et al., 2017) However, the disinfection efficacy may be compromised and the standards can be difficult to achieve in actual practice for several reasons. First, *N. fowleri* associated with pipe wall biofilm has a greater chlorine resistant capability.(Miller et al., 2015) Second, long-distance drinking water piping can lead to reduced chlorine levels, which allows *N. fowleri* to survive and proliferate within water distribution pipelines.(Malinowski et al., 2024; Miller et al., 2015; Momba et al., 2000; Morgan et al., 2016; Pélandakis et al., 2000) In all cases, the occurrence of *N. fowleri* poses a challenge to the water authorities and public safety. Consequently, there is a pressing need to develop rapid diagnostics that enable expedient action by water authorities to minimize public exposure to *N. fowleri*.

The major drawbacks of current cultured-based *N. fowleri* detection methods are the time-consuming and labor-intensive characteristics. On the other hand, the qPCR-based method developed in recent years lacks the capability to indicate *N. fowler* viability in water samples. To supplement the current detection methods, in our previous studies, we developed an untargeted metabolomics method for rapid *N. fowleri* detection based on lab-cultured and field collected samples, respectively. In the lab-cultured sample based effort,(Yu et al., 2017) pathogenic *N. fowleri* and non-pathogenic *N. lovaniensis* and *N. italica* were grown on *E. coli* in separate culture flasks. Untargeted metabolomics analysis revealed that pathogenic *N. fowleri* had different metabolite profiles from non-pathogenic *N. lovaniensis* and *N. italica*. Particularly, 63 metabolite features exhibited significant abundance changes across different species which established the foundation for further development of metabolite-based *N. fowleri* surveillance tools. In the subsequent study, our investigation switched from the strictly controlled lab-cultured setting to field samples.(Yu et al., 2018) In that effort, the field samples (*N. fowleri* positive/negative, *N. lovaniensis* positive) were collected from different sites along two different DWDS systems at several time points from year 2014-2015. Close inspection of the metabolite profiles revealed that 60 significant features consistently differentiated the *N. fowleri* positive from *N. fowleri* negative and *N. lovaniensis* positive filed samples, 10 of which were found to be among the 63 significant metabolite features in the lab-cultured samples study as well. The prediction model built upon the 10 common significant features demonstrated satisfying accuracy when tested on the samples with unknown classification labels.

To complement and extend the previous untargeted metabolomic field study which focused on the comparisons across different locations and DWDS systems, the current effort details the results coming from a year-long (Jan 2017 – Jan 2018) time series study conducted on a single field site known to be seasonally colonized by *N. fowleri*.(Miller et al., 2017) Triplicate biofilm samples were harvested monthly for 13 months, from a biofilm monitor directly connected to the operational drinking water distribution pipeline. In total, 39 biofilm samples reflecting the seasonal changes of *N. fowleri* colonisation at a field site throughout a year were collected and investigated by the untargeted metabolomics approach. The aim of current study is to further validate and improve the previous findings from a temporal perspective. The significant features found in the current study were correlated with results from the prior field survey and their corresponding chemical identities were, when possible, confirmed. In addition, a prediction model for *N. fowleri* surveillance was built upon the seasonal data and was further compared with the model generated in our previous field survey. Relatively high prediction accuracy was obtained from both models when tested on the independent sample data.

## 2. Materials and Methods

### 2.1 Field sample collection and preparation

#### Operational DWDS Field Site

Biofilm samples collected from a KIWA biofilm monitor directly connected to a rural chlorinated regional pipeline in Western Australia (WA) were used to develop the metabolite detection method. This site (SK) was selected due to historical detections of viable free living amoebae (FLA), including *N. fowleri, N. lovaniensis*, and *V. vermiformis* as well as a typically low free chlorine residual (less than 0.1 mg/L).(Miller et al., 2017; Puzon et al., 2009) A biomonitor (KIWA, Netherlands), previously described,(van der Kooij et al., 1995) was connected directly to the DWDSs with a flow rate of 50 L/h and contained up to 42 glass rings, each with a surface area of 16.96 cm^2^, which acted as growth substrates for biofilm formation. Glass was selected as a lower colonizable surface versus other materials, as has been previously reported for laboratory and field based studies.(van der Kooij et al., 1995) Prior to analysis, samples were stored and transported at room temperature and all efforts were made to expediently extract metabolites and conduct amoebae viability tests.

#### Biofilm Samples

Biofilms were sampled and analyzed monthly for one year for the presence of viable and nonviable thermophilic and mesophilic FLA, microbial cell counts, free and total chlorine residuals and water temperature as previously described.(Miller et al., 2017; Morgan et al., 2016; Puzon et al., 2009) For biofilm sampling, triplicate glass rings were removed from the biomonitors and placed in 30 mL of filtered (0.22 µm) site water in a sterile 50 mL tube. Glass rings were vortexed (30 s) and sonicated (5 min, 30 W with a working frequency of 47 kHz + 6% (Bransonic, U.S.A.) to detach the biofilm and cells were harvested by centrifugation at 5000 × g for 10 min. Cell pellets were resuspended in 3 mL of the original solution and the cell concentrate was used for viability plating, flow cytometry, along with DNA and metabolite extractions.

### 2.2 Microbial enumeration and field site physical and chemical parameters

#### Physical and Chemical Measurements

A pocket colorimeter II (Hach, U.S.A.) was used to measure free and total chlorine concentrations, according to the manufacturer’s protocol. Water temperature was measured using the MC-87 Dual Channel Digital Thermometer (TPS, Australia). Turbidity was measured using an Orion AQ4500 turbidity meter (Thermo Scientific, U.S.).

#### Microbial Enumeration

Enumeration of total microbial cell concentrations in biofilm samples were conducted as previously described using a BD Accuri C6 Plus flow cytometer (BD Biosciences, U.S.A.).(Miller et al., 2018a, 2018b) Total cell counts were enumerated following staining with SYBR Green 1 (Invitrogen, U.S.A.). Briefly, before counting in the flow cytometry, the biofilm samples were diluted with MilliQ water to fit into the counting range. A 200 μL aliquot of the diluted sample was stained using 2 μL (10×) of SYBR Green 1 and incubated for 15 min in the dark. Controls included; filtered and/or unstained field samples or Milli-Q water, and flow check beads (CS&T RUO Beads (BD Biosciences, U.S.A.)). All samples were run in triplicate and average values were reported. Results for flow cytometry were calculated as cells/cm^2^.

#### Viability Assessment

Following the protocols outlined in the previous effort, the viability of *Naegleria* was assessed by incubating cell concentrates on non-nutrient agar (NNA)-*E. coli* plates for at least 48 h at 42 °C or 30 °C and observed for plaques.^14^ Plaques were scraped using sterile disposable 1 μL loops and collected for DNA extraction and quantitative polymerase chain reaction (qPCR) for species identification. NNA was prepared by mixing 1 L of 25 % Ringers solution with 15 g of bacteriological agar (Agar No. 1, Oxoid England) and autoclaved at 121 °C for 20 min before plating 100 μL of *E. coli* culture (5.39 × 10^8^ cells/ mL) on the plates. Culturing methods used for laboratory *E. coli* were as previously described.(Miller et al., 2017; Puzon et al., 2009)

#### DNA Extraction

DNA was extracted from biofilm samples using the following two methods. First, PowerSoil DNA Isolation kit (MO BIO Laboratories, U.S.A.) was used for extracting total DNA from cell pellets harvested at 21,000 × g for 5 min from cell concentrates according to the manufacturer’s protocol. Second, the Bio-Rad InstaGene Matrix (Bio-Rad, U.S.A.) was used according to the manufacturer’s protocol for extracting DNA from viable plaques as previously described.(Miller et al., 2018b) Positive NNA-*E. coli* plates were scraped using a 1 μL sterile disposable loop and resuspended in 100 μL of InstaGene matrix. All DNA extracts were stored at −20 °C until analyzed by qPCR.

#### FLA Identification

Methods for FLA identification by qPCR melt curve analysis have previously been described.(Miller et al., 2017; Morgan et al., 2016; Puzon et al., 2017, 2009) In this method, DNA samples were analyzed using a Bio-Rad iQ5 (Bio-Rad, U.S.A.) with a total reaction volume of 25 μL; containing 12.5 μL HotStar Taq Master Mix (2×) (Qiagen, U.S.), 1.25 μL of each primer (10 μM), 0.1 μL 500 μM SYTO9 dye (Molecular Probes, U.S.A.), 7.9 μL sterile double distilled water, and 2 μL of template DNA. Samples were run in triplicate with an *N. fowleri* specific primer set, which only amplifies the intragenic spacer region (ITS) and 5.8S rDNA of *N. fowleri*.^11^ Positive controls (target DNA), DNA extraction method controls (Instagene Matrix or PowerSoil elution buffer with no DNA template) and negative controls (RNase-free H_2_O) were run with every PCR reaction. Positive *N. fowleri* samples were enumerated as total *N. fowleri* cells/cm^2^ by qPCR using standard curves as previously reported.(Miller et al., 2018b; Puzon et al., 2009)

### 2.3 Metabolite Extraction

The metabolite extraction from field samples was performed using a hot methanol approach.(Yu et al., 2017) This method used a quenching solution (60 % methanol vol/vol with 0.85 % AMBIC) that was made by combining 30 mL methanol (VWR Chemicals, USA), 5 mL of 8.5 % (wt/vol) ammonium bicarbonate (AMBIC) (Sigma, USA), and 14.5 mL of MilliQ water. AMBIC 8.5 % was created by dissolving 0.85 g of AMBIC in 10 mL of MilliQ water, adjusted to pH 7.4 with 12 M HCl (Rowe Scientific, Australia) and brought up to a final volume of 50 mL with MilliQ water. Prior to gentle mixing and centrifuging at 5,000 g for 2 min, the samples were quenched using equal volumes of the quenching solution. The supernatant from each sample was subsequently decanted and cell metabolites were extracted from the cell pellet using the hot methanol method. Each cell pellet was resuspended in 500 μL 100 % methanol (pre-heated to 70 °C for 30 min). Samples were then incubated at 70 °C for 15 min. An equal volume (500 μL) of Milli-Q water was added to each replicate and vortexed. The sample was then centrifuged at 12,000 × g for 1 min and the supernatant transferred to a fresh tube. This saved supernatant was then stored at - 80 °C and used for metabolite analysis.

### 2.4 Liquid Chromatography-Quadrupole Time of Flight Analysis

Untargeted metabolite measurements including liquid chromatography (LC)-based metabolite separation, MS analysis, and tandem mass fragmentation were conducted on an ultra-performance liquid chromatography (UPLC) system (Acquity I-Class UPLC, Waters, Milford, MA) coupled with a quadrupole time of flight mass spectrometer (QTOF, Waters Xevo G2, Waters, Manchester, UK).(Yu et al., 2018, 2017) The LC-based metabolite separation was completed using a C18 BEH column (50 × 2.1 mm, 1.7 μm, Waters, Milford, MA) held at 40 °C. LC/MS grade water with 0.1% formic acid (A) and LC/MS grade acetonitrile with 0.1% formic acid (B) were used as solvents at a flow rate of 0.3 mL/min. The gradient method followed the setting in the previous paper for the convenience of data comparison between studies and was as follows: 0-0.2 min: 5% mobile phase B, 0.2 to 2.5 min: increased mobile phase B to 100% linearly, 2.5 to 4 min: 100% mobile phase B, 4 to 4.1 min: decreased mobile phase B to 5%, 4.1 to 5 min: held at 5% for column re-equilibration. The sequence of sample injection was randomized prior to initiating each experimental run. 8 μL of each sample was injected and 4 technical replicates were performed for each sample. A quality control (QC) sample was made by pooling 20 μL from each analytical sample and inserted into the injection queue with an interval of every 12th analytical sample injection to monitor system performance.

The MS data was acquired with positive mode using MS^E^ function (centroid mode, scan time: 0.1 s, low energy: off; high energy: ramp from 20 to 50 eV) over the *m/z* range of 50-1200. The cone gas and desolvation gas was set at a flow rate of 50 and 1000 L/h, respectively. Additionally, the cone and capillary voltage was kept at 35 V and 2.8 kV, respectively. Mass measurement accuracy was calibrated using the lockspray system where leucine enkephalin (2 ng/μL) with *m/z* of 556.2771 was infused every 45 s during data acquisition.

### 2.5 Data Processing and Reduction

Progenesis QI (Non-Linear Dynamics, Durham, NC) software was used for the raw data extraction in which the peak alignment, peak deconvolution, peak picking, and normalization were performed. The *m/z* values, retention times, and corresponding abundance information was then imported into Metaboanalyst for subsequent statistical analysis.(Xia et al., 2009) No data normalization or transformation was performed prior to orthogonal partial least squares discriminant analysis (OPLS-DA), t-test, and fold change calculation in Metaboanalyst since such processes were been completed in Progenesis QI.

### 2.6 Significant feature extraction and identification

The significant features were chosen according to the following criteria: fold change (*N. fowleri* positive vs *N. fowleri* negative) > 1.5, p value (false discovery rate adjusted) < 0.05. The selected significant features with reasonable abundance and positions in S plot of OPLS-DA subsequently were selected for targeted MS fragmentation with the aim of feature identification. For these secondary analysis steps, the scan time of the quadrupole was set at 0.5 s and fragmented collision energy ramped from 5 to 60 eV. The collected MS/MS spectra were screened against a range of metabolic mass spectral databases including Massbank,(Horai et al., 2010) METLIN,(Smith et al., 2005) mzCloud, and HMDB.(Wishart et al., 2012) When possible primary chemical standards were employed to confirm the identity of candidate metabolite signatures. Using the identification guidelines proposed by the Chemical Analysis Working Group (CAWG) MSI (Sumner et al. 2007), the confidence level of the putatively identified metabolites in this effort was equal to 2 where the exact mass and fragment information correspond to a verified entry in the previously listed databases.(Sumner et al., 2007) It is important to recognize that many of the metabolite spectral databases do not contain amoeba-specific entries and the possibility for incorrect assignment exists. It was for this reason that multiple database searches were conducted. With respect to the current effort, only 2 spectral databases returned putative identifications with reasonable levels of confidence. For the spectra considered in this effort the database-specific score thresholds for metabolite candidate consideration were as follows: MassBank > 0.7 and Metlin > 70. Using these score thresholds provides sufficient levels of scrutiny to return a set of spectral matches with high levels of confidence in assignments. Cross referencing of identifications was conducted when possible as well as the use of primary chemical standards.

### 2.7 Prediction model building and evaluation

The prediction model was constructed with the aid of the Biomarker analysis module integrated into Metaboanalyst. The master table containing *m/z* values, retention times, and abundance information as mentioned in the ‘Data Processing and Reduction’ section was imported into Biomarker analysis without any transformation or normalization. The features used for modeling building were chosen with the ‘Builder’ tool. A random forest algorithm was used to construct a prediction model based on the features meeting that were differentially observed and significant. The model performance was evaluated by Receiver Operating Characteristic (ROC) plots with 95% confidence band and the prediction accuracy was computed with 100 cross validation steps.(Xia et al., 2013) The performance of the resulting model an be found in the ‘New Sample Prediction’ section with the accuracy generated by the number of correct classifications.

## 3. Results and Discussion

### 3.1. DWDS field site conditions and *Naegleria* species identification

Field biofilm samples were collected on a monthly basis for 13-months from a single site (SK) to account for seasonal variation in the presence and abundance of *N. fowleri* and other amoebae in this section of operational DWDS. Biofilm samples contained viable *N. fowleri*, in the southern hemisphere summer and autumn months (January – May) (Table 1). Total *N. fowleri* cells in viable samples ranged from >1 to 24.8 cells/cm^2^ as calculated by qPCR (Table 1) which had a temperature range of 14 - 40 °C (average temperature 23.9 °C). Thermophilic amoeba (TA) were detected in the winter and early spring months (July-September) (data not shown) with an average water temperature of 17 °C. Viable *N. fowleri* were detected in biofilm samples with an average biofilm density of 3.04 × 10^5^ cells/cm^2^ and 1.06 × 10^4^ cells/cm^2^, respectively. The average biofilm density of all DWDS biofilm samples was approximately 4.99 × 10^4^ cells/cm^2^. The free chlorine residual was below 0.1 mg/L throughout the duration of the study (Table 1).

**Table 1.**
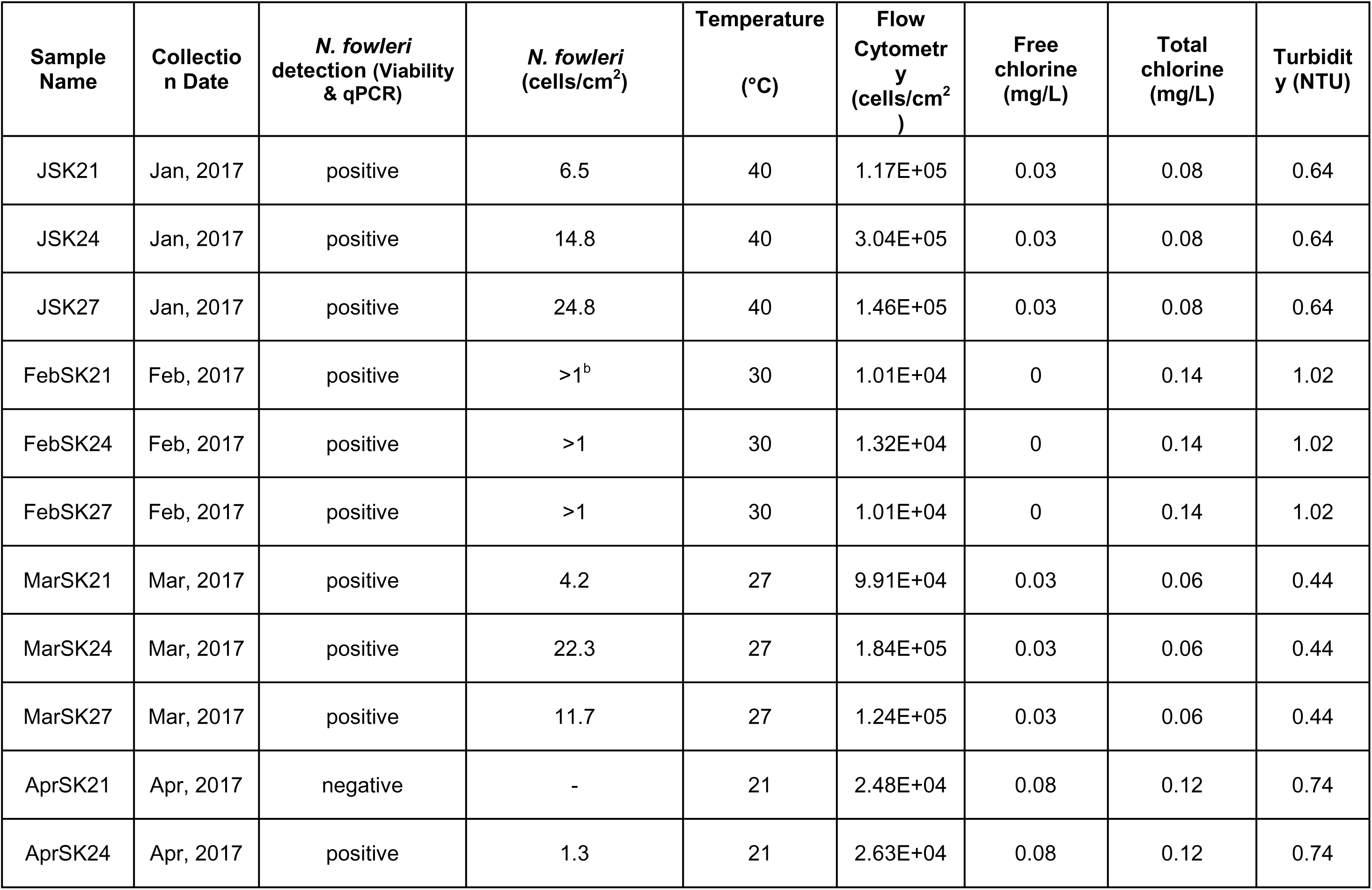
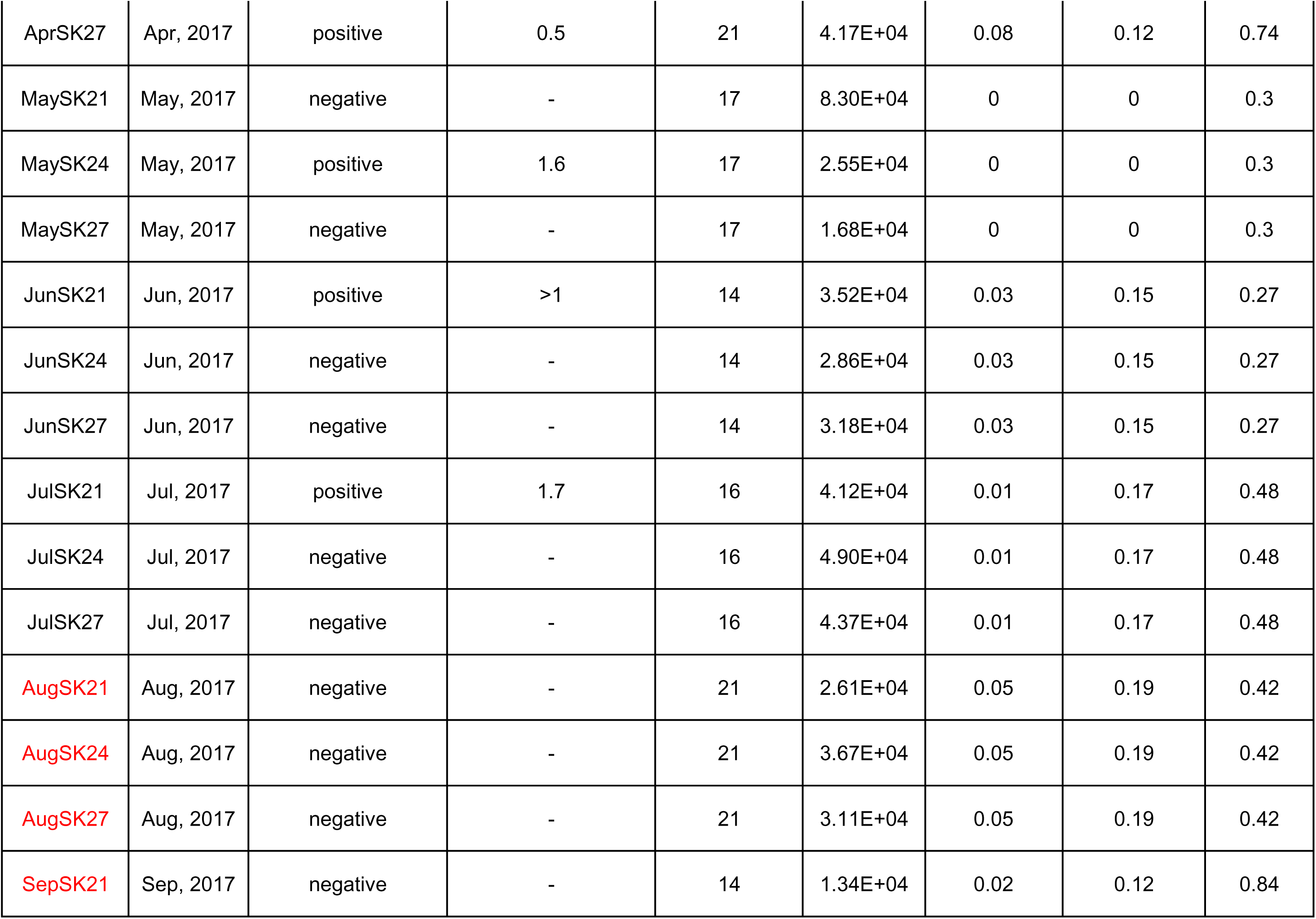
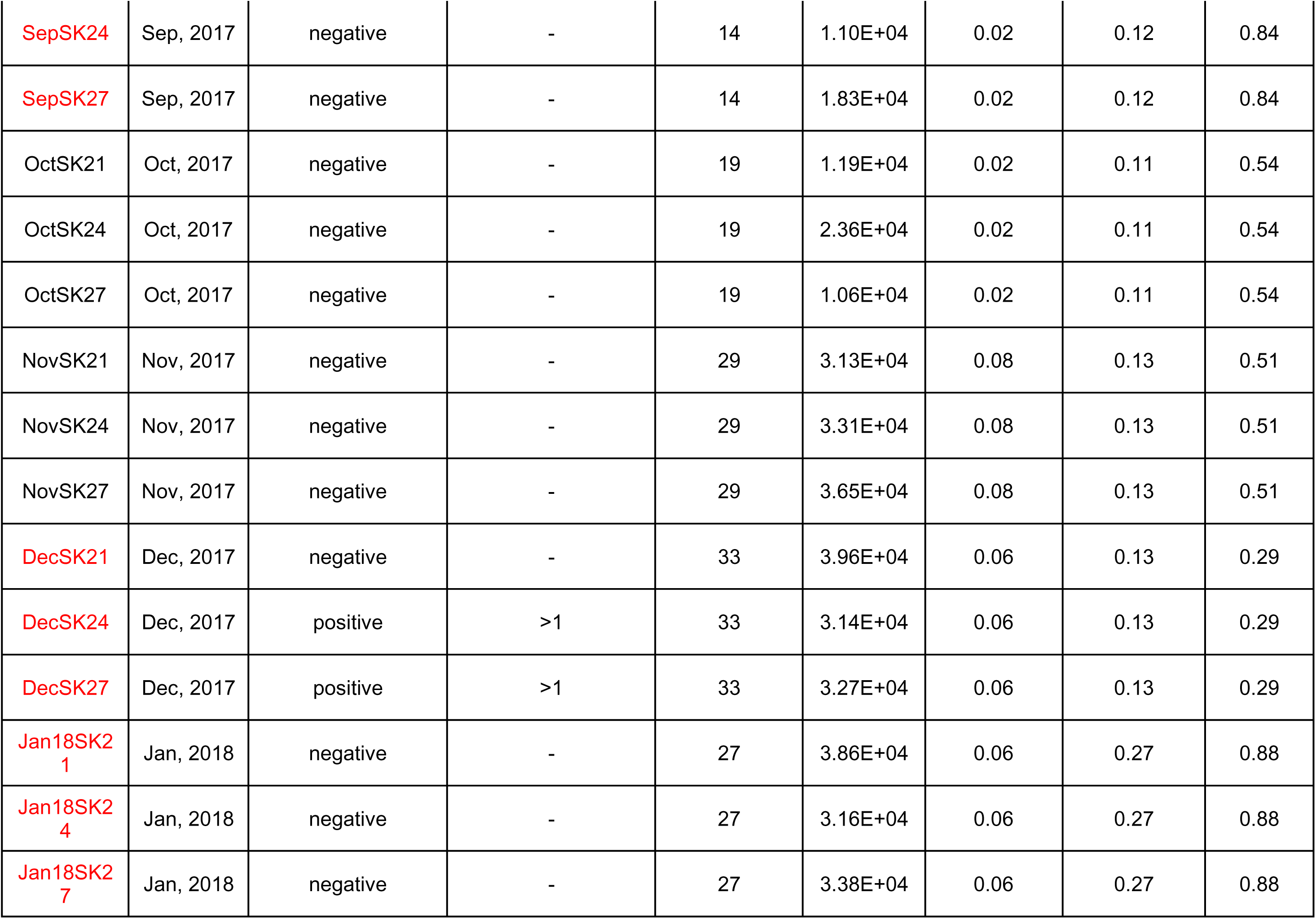
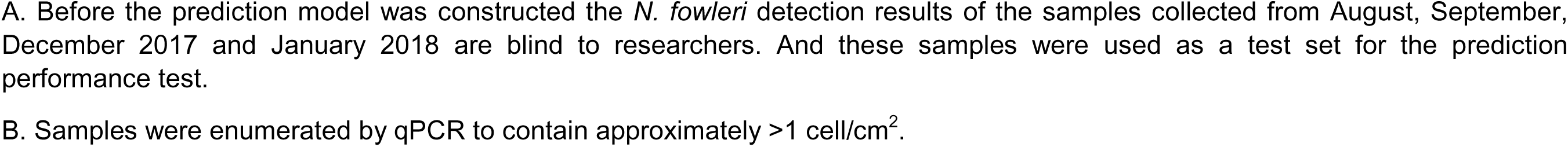
Field sample descriptions.

### 3.2. Multivariate Analyses

Comparison between the *N. fowleri* positive and negative sample groups was performed using OPLS-DA. The *N. fowleri* conditions for the samples collected from August ‘17, September ‘17, December ‘17, and January ‘18 were still unknown during the data analysis and thus excluded in the initial OPLS-DA. Accounting for the 4 technical replicates across each sample yields 108 samples that were, in total, assessed using OPLS-DA. It is worth noting that even though some biological replicate samples were collected at the same time point, their *N. fowleri* conditions (positive or negative) could be different. For example, triplicate biological samples were obtained in April, of which AprSK24 and AprSK27 were positive for *N. fowleri* while AprSK21 was negative (Table 1). Such samples can potentially increase the difficulty of group separation since the samples collected at the same time point are assumed to have similar metabolite profiles though with different group labels. However, the *N. fowleri* positive and negative samples are still clearly separated through the T score axis of the OPLS-DA score plot, as shown in Figure 1. The calculated goodness-of-fit parameters for the OPLS-DA model were: R2X = 0.210, R2Y = 0.785, Q2 = 0.733. We considered the OPLS-DA model to be robust because the R2Y > 50% and Q2 > 50%.(Blasco et al., 2015) The S-plot (Figure S1) was also generated as a reference for the significant features identification in the following section.

**Figure 1.**
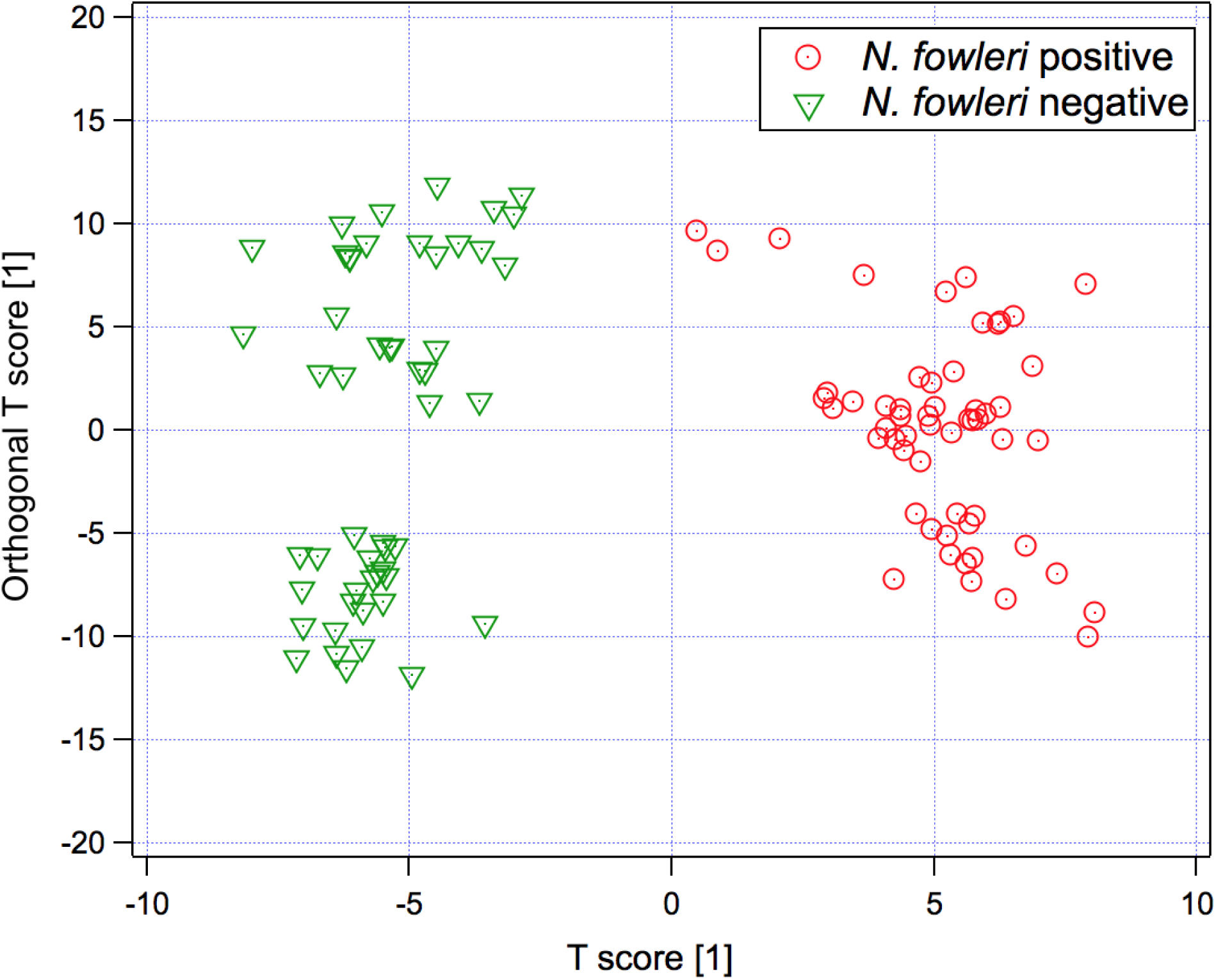
Orthogonal partial least-square discriminant analysis (OPLS-DA) scores plot of *N. fowleri* positive (red dots) vs. negative (green triangles) samples. R2X = 0.210, R2Y = 0.785, Q2 = 0.733.

### 3.3. Significant Feature Identification

To evaluate the significant features that differentiate the *N. fowleri* positive from negative samples, a criteria of p values (FDR adjusted) < 0.05 and fold change (*N. fowleri* positive/*N. fowleri* negative or *N. fowleri* negative/*N. fowleri* positive) > 1.5 was applied. In total, 161 significant features that contribute to *N. fowleri* positive/negative separation were found (Table S1). After collectively considering the significant feature abundances and its corresponding position in the S-plot, 81 out of 161 were selected to proceed to the targeted fragmentation step as described in the experimental section for feature identification. The obtained MS/MS spectra were matched with the available information in online databases. However, as mentioned in our previous papers,(Yu et al., 2018, 2017) the amoebae-specific metabolite information is very limited in the public databases, which hinders the comprehensive metabolite identification of significant features. Finally, the chemical identities of 2 significant features were confirmed and their corresponding information including *m/z*, retention, adduct form, fold change, change direction, and p-value are summarized in Table 2. Their positions in the S plot of OPLS-DA are indicated in Figure S1. As demonstrated in Figure 2, the MS/MS patterns of feature *m/z* 268.1034 and 152.0565 match with that of adenosine and guanine, respectively. The mass measurement accuracy errors are off by 4 (adenosine) and 5 (guanine) ppm, respectively, which is in a normal range of our instrument performance. In addition, both adenosine and guanine displayed markedly upregulated fold changes (adenosine: 1179, guanine: 8) and low p values (adenosine: 1.56×10^-4^, guanine: 2.55×10^-4^) in *N. fowleri* positive/negative comparison (Table 2).

**Figure 2.**
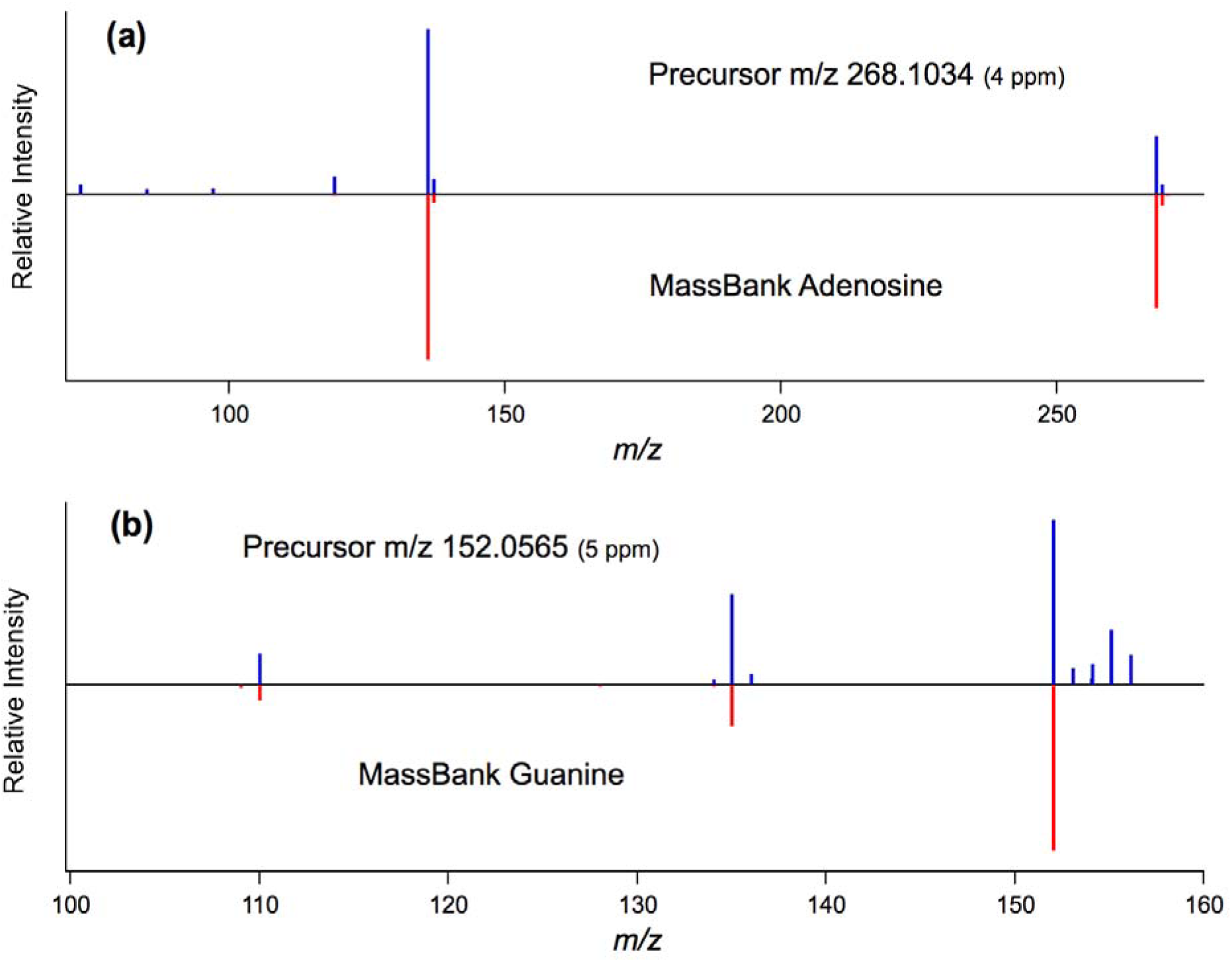
Tandem mass spectra match for identification validation. (a) matching with the MS/MS data of Adenosine in MassBank (b) matching with the MS/MS data of Guanine in MassBank.

**Table 2.** Putative chemical identification, adducts, fold changes and mass accuracy of significant features.

| *m/z* | Retention time (min) | adduct | Mass Error (ppm) | Fold change | up/down regulated (positive vs negative) | p-value (FDR adjusted) | Identification |
| --- | --- | --- | --- | --- | --- | --- | --- |
| 268.1034 | 0.59 | [M+H]<sup>+</sup> | 4 | 1179.2 | up | 1.56×10<sup>-4</sup> | Adenosine |
| 152.0565 | 0.43 | [M+H]<sup>+</sup> | 5 | 8.4 | up | 2.55×10<sup>-4</sup> | Guanine |

The box plots of adenosine and guanine are shown in Figure 3, where adenosine and guanine abundance for the *N. fowleri* negative samples were close to 0 while the *N. fowleri* positive samples have wide abundance range for adenosine and guanine. Figure 4 revealed more details of the adenosine and guanine abundance across months. The monthly metabolite levels are further stratified based on the different biological triplicates (sample #21, #24, #27) and the *N. fowleri* condition for each month is indicated by background color where light red represents *N. fowleri* positive and light blue represents *N. fowleri* negative. In Figure 4, the adenosine and guanine abundance in *N. fowleri* negative months are generally at a low level for all the triplicate biological samples. However, the metabolite abundance of *N. fowleri* positive samples can vary remarkably depending on the months and specific biological triplicate. For example, sample #24 collected from Jan 2017 to May 2017 are all *N. fowleri* positive. Close inspection indicates that the adenosine abundance was above 2.0 × 10^4^ in the months such as January, March, and May. But the adenosine level was lower than the detection limit in February and April. Similar behavior can also be observed in the guanine data. In sample #27, relatively high guanine levels could be detected in the *N. fowleri* positive months, February and April. But in the other two *N. fowleri* positive months, January and March, guanine level was significantly lower.

**Figure 3.**
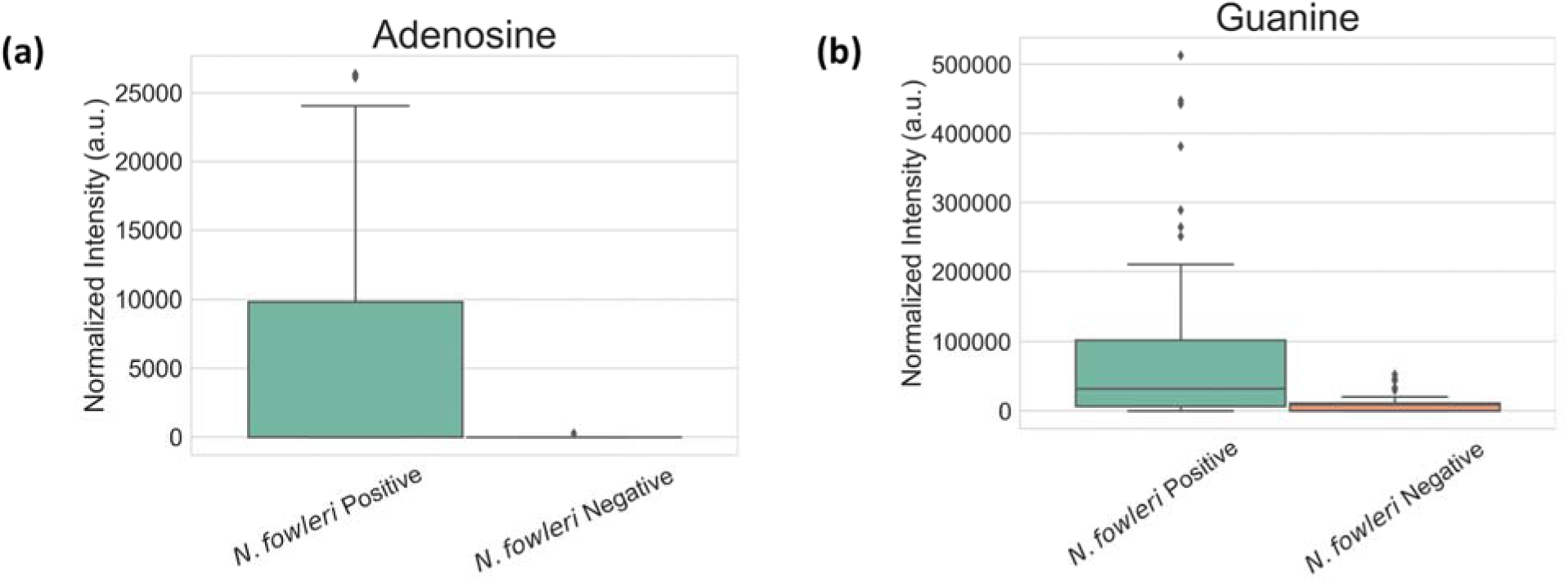
Box plot profiles (N. f. positive vs. N. f. negative) for significant metabolites. (a) Adenosine (b) Guanine.

**Figure 4.**
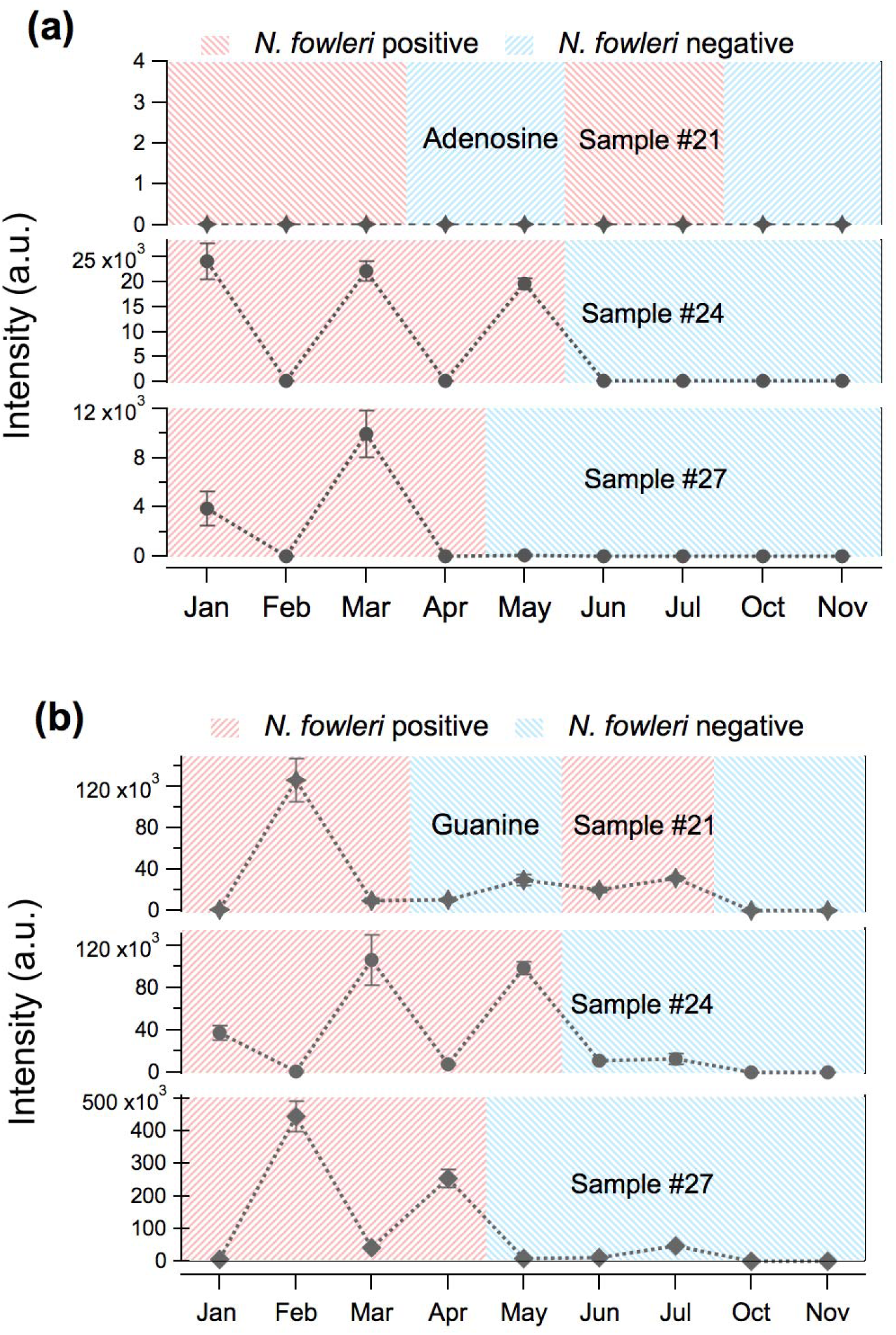
The abundance changes of (a) adenosine and (b) guanine in biological triplicates samples #21, #24, and #27 throughout a year.

Collectively considering the data presented in Figure 3 and 4, it can be preliminarily concluded that although all the *N. fowleri* negative samples have low adenosine and guanine abundance, the low adenosine and guanine abundance does not necessarily imply *N. fowleri* negative condition because some of the *N. fowleri* positive samples also have low adenosine and guanine levels. However, if the intensities of adenosine and guanine detected are above a certain level, it stands to reason that a warning (and proactive DWDS management strategies) should be raised that *N. fowleri* may appear in the samples because *N. fowleri* negative samples generally exhibit a low adenosine and guanine level in our observed data. There are several possible reasons for the highly fluctuating metabolite intensities in the *N. fowleri* positive samples. The first possible reason is that the interference from other amoebae in the environment. Beside *N. fowleri*, the other amoebae such as *Vermamoeba* and thermophilic amoebae are also detected in the samples, and the appearance of other amoebae can potentially affect the *N. fowleri* metabolite profile. *Miller et al.* investigated the interactions between *N. fowleri* and other free-living amoebae (FLA) such as *Vermamoeba*, *Willaertia*, and *Vahlkampfia* spp. in biofilms, and found that competition existed in the cocolonization of *N. fowleri* and FLA possibly owing to the direct food sources competition.^13,14^ In addition, the seasonal changes can impact the environmental temperature, disinfectant residuals, and nutrient level. All the factors mentioned above can change the *N. fowleri* metabolite profiles which leads to variations of adenosine and guanine levels. Also, it is worth noting that the triplicate biological samples were collected from three independent glass rings in the field site. Even though the biological triplicates were collected at the same time, the *N. fowleri* population was likely to be unevenly distributed in the glass rings which partially explains why adenosine and guanine levels were different among the samples. Additional work is still needed to investigate the reasons for the adenosine and guanine level fluctuation and determine a reasonable warning threshold of adenosine and guanine for *N. fowleri* appearance.

When taking a look back at the identified features in our previous studies, it is interesting to note that adenosine and guanine were also found and identified in the field survey-based and lab-cultured based studies. Similar to the trend observed in the current seasonal study, guanine has a much higher level in the *N. fowleri* positive samples than in the culture media (*E. coli*) and non-pathogenic *N. lovaniensis* and *N. italica* samples. As stated in our previous paper, our rationale for the high guanine level is that *N. fowleri* may involve into a more active proliferation activity but further study is still required to confirm its biological role in *N. fowleri*. In our last field survey-based study,(Yu et al., 2018) there was an ambiguous identification of the significantly changed feature *m/z* 136.060 to be adenine or an adenosine fragment. The MS/MS pattern of *m/z* 136.060 matched perfectly with that of adenine. However, it is worth noting that adenine is also a component of the larger adenosine structure, so possibility remains that the *m/z* 136.060 signal was derived from in-source fragmentation of adenosine. In the current study, both *m/z* 268.1034 and 136.060 with the same retention time of 0.59 min were found and the targeted MS/MS of *m/z* 268.1034 agreed with that of adenosine in database, which proved that the experimental conditions used in the previous and current studies can generate an adenine fragment from adenosine which is understandable as adenosine is composed of adenine connected to ribose sugar. As an important building block of the energy transfer molecules, adenosine triphosphate (ATP) and adenosine diphosphate (ADP), the increased level of adenosine may be related to enhanced energy transfer cycle, which may contribute to the proliferation activity. Recently, the crystal structure of S-adenosyl-L-homocysteine hydrolase from *N. fowleri*, an enzyme catalyzing adenosine synthesis, was deposited into Protein Data Bank.((ssgcid) and Seattle Structural Genomics Center for Infectious Disease (SSGCID), 2017) Interestingly, this enzyme extracted from *N. fowleri* is bound with nicotinamide adenine dinucleotide (NAD) and adenosine in its crystal structure, showing the details of adenosine synthesis in *N. fowleri*.

### 3.4. Prediction model building and comparison

A total of 161 significant features were identified that distinguish *N. fowleri* positive samples from *N. fowleri* conditions. When correlating the 161 significant features with the significant features found at the SK site in our previous field survey-based study, 29 common features (i.e. mass difference < 20 ppm, retention time difference < 0.05 min) were found to play an important role in sample differentiation in both previous and current studies. The corresponding pertinent characteristics of the 29 common features is summarized in Table S2. In addition to the SK site, an additional sample set was included in the current effort from the KT site. In Table S3* of our prior field survey-based manuscript,(Yu et al., 2018) the common significant features that appeared in both SK and KT sites were listed. Beyond just correlating the SK site significant features, we performed an addition further analysis by correlating the current results from seasonal measurements at SK with the survey measurements at SK and KT (Table S3*). This additional distillation of features using both SK & KT sites results in a more restrictive measure for including a feature in the predictive model. The consequence of this additional comparison is a reduction in the number of entries in Table S2 from 29 to 24. This final list of 24 is shown in Supplementary Materials Table S3.

The list consisting of 24 common features was considered to be a robust list of features confidently indicating the presence of viable *N. fowleri* while excluding and was further used to build a prediction model in the next step. Before model building, 4 out of 24 features with retention time < 0.40 min were excluded since their retention time was close to the column’s void volume retention time, indicating poor retention on the column. Finally, a prediction model was built using the 20 common features with the random forest algorithm and the aid of the Biomarker analysis module in Metaboanalyst. The model performance was evaluated by ROC curve analysis with the result demonstrated in Figure 5. As shown in Figure 5, the prediction model achieved a relatively high area under curve (AUC) score of 0.946 with a confidence interval of 0.843-1, representing a relatively high model sensitivity and specificity. The confusion matrix of the prediction model is shown in Table S1. ‘0’ in the table represents ‘*N. fowleri* negative’ and ‘1’ represents ‘*N. fowleri* positive’. Closer examination of the table illustrates that based upon the predictive model using the significant features previously described ten *N. folweri* negative samples were misclassified as positive while two *N. fowleri* negative samples were misclassified as being positive. According to the confusion matrix, the false positive rate (FPR) and false negative rate (FNR) of the prediction model were calculated as 19.2% and 3.6%, respectively. To further confirm the prediction model performance, the samples collected from August ‘17, September ‘17, December ‘17, and January ‘18 were used as an independent test set. It is worth noting that the *N. fowleri* conditions for these samples were unknown to researchers until the prediction model was built and prediction results were generated. The prediction results are shown in Table 3. Inspection of Table 3 reveals that the predictions from our newly established seasonal prediction model generally have high confidence levels, with the probability in the range of 0.880 – 0.980. Comparing the prediction results with the actual *N. fowleri* conditions in Table 1, we found that the seasonal prediction model has a satisfying accuracy of 83.3% with only the samples DecSK24 and DecSK27 were incorrectly labeled. The prediction for samples DecSK21, DecSK24, and DecSK27 can be particularly challenging. These three samples collected at the same time in December 2017 which were expected to have similar metabolite profiles, turned out to have different *N. fowleri* conditions where DecSK21 was negative for *N. fowleri* and DecSK24 and DecSK27 were positive. The prediction results imply that the current metabolite panel used for model building requires additional refinement to reduce the FPR. However, in its current state the prediction model FNR is < 5% which may prove useful when assessing the water quality.

**Figure 5.**
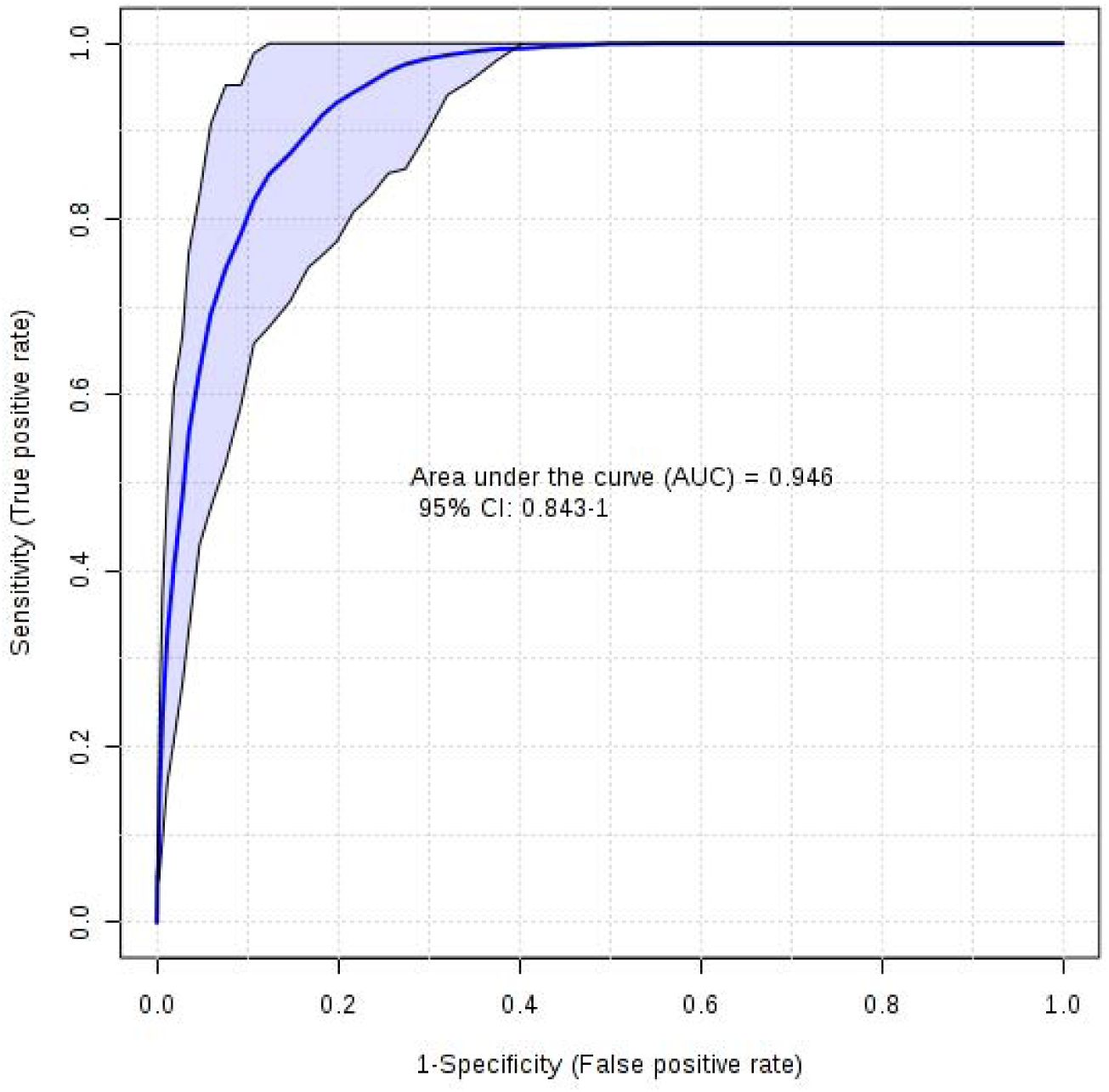
The ROC curve analysis for the prediction model built on the significant common features with the previous field sample study. Model was built on a random forest algorithm with AUC of 0.946 and CI of 0.843-1 for distinguishing *N. fowleri* positive from *N. fowleri* negative samples.

**Table 3.** Prediction results comparison between the seasonal and site survey models.

| Sample name | Seasonal Model |  | Site Survey Model |  |
| --- | --- | --- | --- | --- |
|  | Probability | Class | Probability | Class |
| AugSK21_A | 0.897 | *N. fowleri* negative | 1.00 | *N. fowleri* negative |
| AugSK21_B | 0.917 | *N. fowleri* negative | 1.00 | *N. fowleri* negative |
| AugSK21_C | 0.907 | *N. fowleri* negative | 1.00 | *N. fowleri* negative |
| AugSK21_D | 0.897 | *N. fowleri* negative | 1.00 | *N. fowleri* negative |
| AugSK24_A | 0.890 | *N. fowleri* negative | 1.00 | *N. fowleri* negative |
| AugSK24_B | 0.903 | *N. fowleri* negative | 1.00 | *N. fowleri* negative |
| AugSK24_C | 0.927 | *N. fowleri* negative | 1.00 | *N. fowleri* negative |
| AugSK24_D | 0.920 | *N. fowleri* negative | 1.00 | *N. fowleri* negative |
| AugSK27_A | 0.893 | *N. fowleri* negative | 1.00 | *N. fowleri* negative |
| AugSK27_B | 0.907 | *N. fowleri* negative | 1.00 | *N. fowleri* negative |
| AugSK27_C | 0.913 | *N. fowleri* negative | 1.00 | *N. fowleri* negative |
| AugSK27_D | 0.910 | *N. fowleri* negative | 1.00 | *N. fowleri* negative |
| SepSK21_A | 0.980 | *N. fowleri* negative | 1.00 | *N. fowleri* negative |
| SepSK21_B | 0.950 | *N. fowleri* negative | 1.00 | *N. fowleri* negative |
| SepSK21_C | 0.960 | *N. fowleri* negative | 1.00 | *N. fowleri* negative |
| SepSK21_D | 0.953 | *N. fowleri* negative | 1.00 | *N. fowleri* negative |
| SepSK24_A | 0.953 | *N. fowleri* negative | 1.00 | *N. fowleri* negative |
| SepSK24_B | 0.950 | *N. fowleri* negative | 1.00 | *N. fowleri* negative |
| SepSK24_C | 0.947 | *N. fowleri* negative | 1.00 | *N. fowleri* negative |
| SepSK24_D | 0.937 | *N. fowleri* negative | 1.00 | *N. fowleri* negative |
| SepSK27_A | 0.940 | *N. fowleri* negative | 1.00 | *N. fowleri* negative |
| SepSK27_B | 0.957 | *N. fowleri* negative | 1.00 | *N. fowleri* negative |
| SepSK27_C | 0.947 | *N. fowleri* negative | 1.00 | *N. fowleri* negative |
| SepSK27_D | 0.963 | *N. fowleri* negative | 1.00 | *N. fowleri* negative |
| DecSK21_A | 0.900 | *N. fowleri* negative | 1.00 | *N. fowleri* negative |
| DecSK21_B | 0.917 | *N. fowleri* negative | 1.00 | *N. fowleri* negative |
| DecSK21_C | 0.903 | *N. fowleri* negative | 1.00 | *N. fowleri* negative |
| DecSK21_D | 0.963 | *N. fowleri* negative | 1.00 | *N. fowleri* negative |
| DecSK24_A | 0.900 | *N. fowleri* negative | 1.00 | *N. fowleri* negative |
| DecSK24_B | 0.930 | *N. fowleri* negative | 1.00 | *N. fowleri* negative |
| DecSK24_C | 0.917 | *N. fowleri* negative | 1.00 | *N. fowleri* negative |
| DecSK24_D | 0.920 | *N. fowleri* negative | 1.00 | *N. fowleri* negative |
| DecSK27_A | 0.913 | *N. fowleri* negative | 1.00 | *N. fowleri* negative |
| DecSK27_B | 0.967 | *N. fowleri* negative | 1.00 | *N. fowleri* negative |
| DecSK27_C | 0.937 | *N. fowleri* negative | 1.00 | *N. fowleri* negative |
| DecSK27_D | 0.923 | *N. fowleri* negative | 1.00 | *N. fowleri* negative |
| Jan18SK21_A | 0.970 | *N. fowleri* negative | 1.00 | *N. fowleri* negative |
| Jan18SK21_B | 0.967 | *N. fowleri* negative | 1.00 | *N. fowleri* negative |
| Jan18SK21_C | 0.957 | *N. fowleri* negative | 1.00 | *N. fowleri* negative |
| Jan18SK21_D | 0.973 | *N. fowleri* negative | 1.00 | *N. fowleri* negative |
| Jan18SK24_A | 0.920 | *N. fowleri* negative | 1.00 | *N. fowleri* negative |
| Jan18SK24_B | 0.893 | *N. fowleri* negative | 1.00 | *N. fowleri* negative |
| Jan18SK24_C | 0.977 | *N. fowleri* negative | 1.00 | *N. fowleri* negative |
| Jan18SK24_D | 0.907 | *N. fowleri* negative | 1.00 | *N. fowleri* negative |
| Jan18SK27_A | 0.947 | *N. fowleri* negative | 1.00 | *N. fowleri* negative |
| Jan18SK27_B | 0.937 | *N. fowleri* negative | 1.00 | *N. fowleri* negative |
| Jan18SK27_C | 0.910 | *N. fowleri* negative | 1.00 | *N. fowleri* negative |
| Jan18SK27_D | 0.880 | *N. fowleri* negative | 1.00 | *N. fowleri* negative |

Next, we evaluated the site survey prediction model from our previous field-sample based study with the test data set (August ‘17, September ‘17, December ‘17, and January ‘18 sample data) and compared its performance with the newly established seasonal prediction model described above. The site survey model was based upon 10 selected significant features found in our previous study. The predictions generated from the site survey model can also be found in Table 3. Cross comparison was conducted between the predictions from the seasonal model and the site survey model. Surprisingly, the class labels predicted by the seasonal and site survey models for the test samples were the same, meaning that the site survey model also had a high degree of accuracy for the current sample set (83.3%). The site survey model also failed in sample DecSK24 and DecSK27 prediction, reiterating the challenges posed by samples with similar metabolite profiles but different *N. fowleri* conditions. This may also reflect a subtle contribution of the supporting biofilm microbial ecology and the abundance of *N. fowleri* food sources to the metabolite profiles.(Miller et al., 2018a; Morgan et al., 2016; Puzon et al., 2017) Though the site survey prediction model came from the biofilm samples collected in 2014-2015, it was still effective in predicting the *N. fowleri* conditions of 2017-2018 samples in the current study. The most significant difference between the prediction results coming from the site survey and the seasonal model is that the prediction probability of the site survey model approached 1, meaning that the model had a higher confidence for its predictions even if two samples, DecSK24 and DecSK27, were misclassified. Also, it is interesting to note that, of the 10 features comprising the site survey model, 4 still displayed significant changes in the current study. The remaining features from the site survey model did not meet our criteria and were excluded from the significant feature list. Overall, the seasonal and site survey models both displayed similar and satisfying prediction accuracy for the independent test data. While the FNR of the seasonal prediction model was low (3.6%), FPR was elevated (19.2%), which exemplifies the need for continued efforts focusing on the metabolism of *N. fowleri* in DWDSs.

## 4. Conclusion

It is clear that an effective *N. fowleri* monitoring method is needed by water utilities to address *N. fowleri* colonization and maximize public health outcomes. However, the existing *N. fowleri* detection approaches are far from satisfying due to their time-consuming, labor intensive characteristics as well as the inability to assess viability. Based on our previous efforts to develop a rapid *N. fowleri* detection method focusing on the lab-cultured and field-survey samples, this manuscript presents our recent progress on the untargeted metabolomics method development. Using a simplified OPLS-DA approach as a visualization tool (Figure 1), it is clear that the untargeted metabolomics approach can serve as a binary classification tool (i.e. presence vs absence). While subtle changes in metabolite profiles were found following the colonization and seasonal loss cycle of *N. fowleri* at the SK site, more than 160 features displayed significant changes (fold change > 1.5, q value < 0.05) between *N. fowleri* positive vs. negative samples. Of the significant features, the chemical identities of guanine and adenosine were successfully confirmed. Monthly analysis of the guanine and adenosine intensity fluctuation indicated that low metabolite levels in samples do not necessarily represent the *N. fowleri* negative condition while the high metabolite levels, on the other hand, can give a warning of possible presence of *N. fowleri*. Further evaluation and refinement of the respective models (seasonal vs. site survey) yield the statistics related to the accuracy of the respective models and suggest that continued investigation can aid in the development of a rigorous threshold for providing warnings to water quality authorities. When accounting for the common features between the current seasonal survey and the previous site survey a total of 24 common features were identified. The prediction model constructed with these common features exhibited satisfying performance with AUC score of 0.946 and accuracy of 83.3% when tested on the independent sample data whose *N. fowleri* condition remained blind to researchers during model construction. Despite practical limitations to the range and numbers of samples collected, the metabolomics based effort continues to demonstrate the potential to rapidly identify the presence of *N. fowleri* in DWDSs. Because the water origin and distribution systems will not be the same as those evaluated in this study, the metabolomics-based method would, in all likelihood, need to be tailored to each drinking water network. Nevertheless, the initial signatures may serve as the bases for initial characterization. Most importantly, the extension of this approach from a controlled laboratory environment to a seasonal site survey demonstrates an adaptable framework for rapid *N. fowleri* monitoring. The continued development and refinement of untargeted-metabolomics based approaches may prove a rapid and useful tool to aid water utilities and water parks with improved surveillance and monitoring of amoebic and other microbial pathogens in operational DWDSs.

## Data Availability

All data produced in the present work are contained in the manuscript

## Supplementary materials

**Figure S1.**
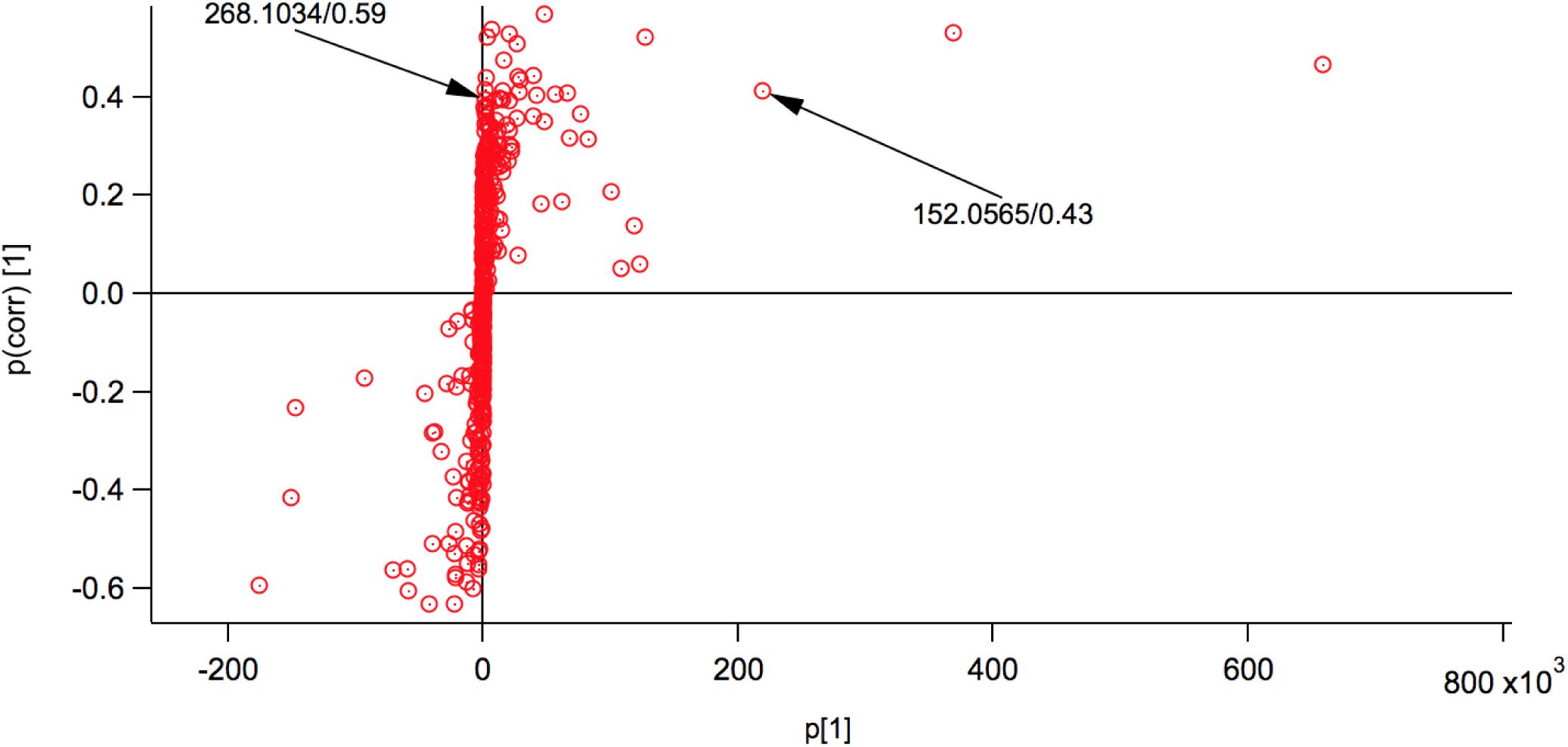
S plot generated from OPLS-DA. The positions of guanine and adenosine are labeled with their *m/z* and retention time.

**Table S1.** Confusion Matrix (cross-validation)

|  | 0 | 1 |
| --- | --- | --- |
| 0 | 42 | 2 |
| 1 | 10 | 54 |

